# The impact of Anxiety, Sleep Quality, Social Media Use, and Socioeconomic Background on Academic Performance in Bangladeshi Public University Students: A Structural Equation Modeling Approach

**DOI:** 10.64898/2026.08.18.26360659

**Authors:** Sagor Ahmad Rana, Md Arafat Hossen, Mohammad Arifur Rahman, Sanjida Tasnim

## Abstract

**Background:** Academic achievement is crucial for university students, but various factors affect their performance. This study explores the impact of anxiety, sleep quality, social media use, and socioeconomic status on academic performance (CGPA) among public university students in Bangladesh.

**Data and Methods:** Data were collected from 225 students using a structured questionnaire that assessed anxiety (GAD-7), sleep quality (PSQI), social media use (SMUQ), and socioeconomic status (income, parental education). Structural Equation Modeling (SEM) was used to analyze the relationships between these variables.

**Outcomes:** The results showed that socioeconomic status had a strong positive effect on academic performance (β = 0.745, p < 0.001), while anxiety negatively impacted academic outcomes (β = - 0.675, p < 0.001). Sleep quality was positively related to academic performance (β = 0.113, p < 0.05), but with a weaker effect. Social media usage is found to have a negative significant effect on academic performance (β = -0.137, p < 0.001).

**Conclusion:** These findings highlight the importance of controlling social media usage and anxiety to enhance academic performance among adult students. Sleep quality and socioeconomic background of the students are also found to be meaningfully associated with their educational progress.

## Introduction

In today’s digitally connected world, social media has become an integral part of young people’s daily lives. Platforms such as Instagram, TikTok, Snapchat, and X (formerly Twitter) offer instant communication, entertainment, and a space for self-expression, but their pervasive use has raised growing concerns about their effects on the well-being of adolescents and young adults [1]. University students, in particular, are among the heaviest users of social media, often spending several hours each day scrolling, posting, and engaging with online content [2]. While these platforms provide opportunities for connection and information sharing, increasing evidence suggests that excessive social media use is associated with users’ mental health [3].

Among the most frequently reported concerns are disturbances in sleep quality and heightened levels of anxiety, particularly social anxiety. Screen time is found associated with shortened duration of sleep and delayed sleep [4]. Late-night screen use, exposure to blue light, and the cognitive and emotional stimulation provided by social media can interfere with sleep onset, duration, and quality. Studies have found consistent, substantial, and progressive associations between social media use and sleep disturbance among young students [5].

Simultaneously, constant exposure to curated online content, social comparison, fear of missing out (FoMO), and cyberbullying contribute significantly to elevated psychological distress among students. Multiple studies have established a consistent negative correlation between FoMO and well-being, showing that higher FoMO is associated with increased anxiety, depression, and a diminished sense of psychological balance [6]. A recent systematic review of 32 studies involving nearly 30,000 university students found that cybervictimization was significantly linked to anxiety in 12 out of 15 studies, with prevalence rates ranging from 27% to 84.1%, and was also associated with decreased academic concentration and productivity [7]. These intertwined issues — disrupted sleep and heightened anxiety — are not isolated problems; they may further undermine students’ academic performance by impairing concentration, memory, motivation, and overall learning outcomes.

Evidence from Asian college populations further reinforces the same concern. In a large cross-sectional study of 4,747 Chinese college students, found that high screen time was significantly associated with anxiety, depression, psychopathological symptoms, and poor sleep quality. Their findings also revealed that low physical activity combined with high screen time interactively elevated the risk of mental health problems and poor sleep, underscoring the compounding effect of sedentary, screen-dominated lifestyles [8]. Similar patterns have been observed across other student populations, where prolonged digital engagement is consistently linked to declining mental health and academic difficulties [9].

Sleep deprivation and anxiety, in particular, are known to impair attention, memory consolidation, and emotional regulation — all of which are foundational to academic success [10]. Recent study shows significant path identified based on indirect effects between anxiety, adaptation problems, academic stress and sleep quality [11]. Poor quality of sleep was significantly associated with elevated mental stress levels [12]. When students engage in heavy social media use, they may experience a reinforcing cycle: disrupted sleep heightens anxiety, while anxiety further disrupts sleep, ultimately diminishing classroom engagement and academic outcomes. This interconnected dynamic is especially pronounced among university students, who already face high academic demands and developmental transitions.

On top of these individual factors, socioeconomic condition remains a significant determinant of academic success. Students from lower socioeconomic backgrounds often face additional barriers such as financial struggles, limited access to academic resources, and increased stress due to low family income [13]. Some students have to financially support their families, and this extra responsibility creates additional pressure that interferes with their studies. Several studies have demonstrated a strong correlation between socioeconomic condition and student academic performance, with students from higher socioeconomic backgrounds generally outperforming their peers from lower economic backgrounds in terms of test scores, grades, and broader educational outcomes [14].

Although several studies have explored anxiety, sleep quality, social media use, and socioeconomic condition separately, very few have examined their combined effect on academic performance. Islam et al. found that Bangladeshi students with high anxiety levels struggled not only with academic tasks but also with maintaining a balance between their personal lives and educational goals [15]. Research by Hossain and Khan indicates that poor sleep patterns among Bangladeshi students are directly linked to poorer academic performance, creating a vicious cycle of exhaustion and declining grades [16]. Socioeconomic status plays a similar role in Bangladesh as it does globally; Islam et al. [15] also reported that students from lower socioeconomic backgrounds often face financial hardships that limit their ability to afford essential academic resources, such as textbooks, online subscriptions, or even reliable internet access [17].

This study aims to address four factors — anxiety, sleep quality, social media use, and socioeconomic condition —are individually interconnected and impact academic performance, for Bangladeshi public university students. By shedding light on the interplay between those factors employing Structural Equation Modeling (SEM), this research tries to capture the behavioral changes encountered by young adults of Bangladesh.

## Methods

### Sample selection and data collection

The study was designed to investigate how anxiety, sleep quality, social media use, and socioeconomic condition interact to impact student academic performance. In this pursuit, 225 respondents were selected from various public universities in Bangladesh, which included both resident and non-resident students from science, business, and humanities disciplines. Data was collected through a questionnaire survey from September 2 to December 15, 2024. The study included full-time students from public universities in Bangladesh who voluntarily participated and provided informed consent prior to data collection.

### Questionnaire Setting

A structured questionnaire was used to gather data from participants (Table 1). The questionnaire was divided into four sections based on the objectives of the study—the first section of the questionnaire assesses the socioeconomic condition of the respondents. The demographic and economic parameters are set according to modified Kuppuswamy scale [18]. Responses were recorded on a Likert scale, with predefined categories for each question. The second section of the questionnaire was designed to estimate respondent’s anxiety level using 7-item Generalized Anxiety Disorder scale (GAD-7) [19]. Each respondent measured on a 5-point Likert scale ranging from 1 (Never) to 5 (Always). The third section, which comprised sleep quality, was adapted from the Pittsburgh Sleep Quality Index (PSQI) [20]. The last section of the questionnaire focuses on social media use, which is designed based on Social Media Use Questionnaire (SMUQ) developed by Xanidis and Brignell [21]. The original SMUQ comprised 21 items; however, following principal component analysis, the instrument was reduced to 9 items representing two dimensions of problematic social networking site use: Withdrawal and Compulsion. In this study, the questionnaire consisted of nine items, each rated on a 5-point Likert scale, ranging from 1 (Never) to 5 (Always).

**Table 1:**
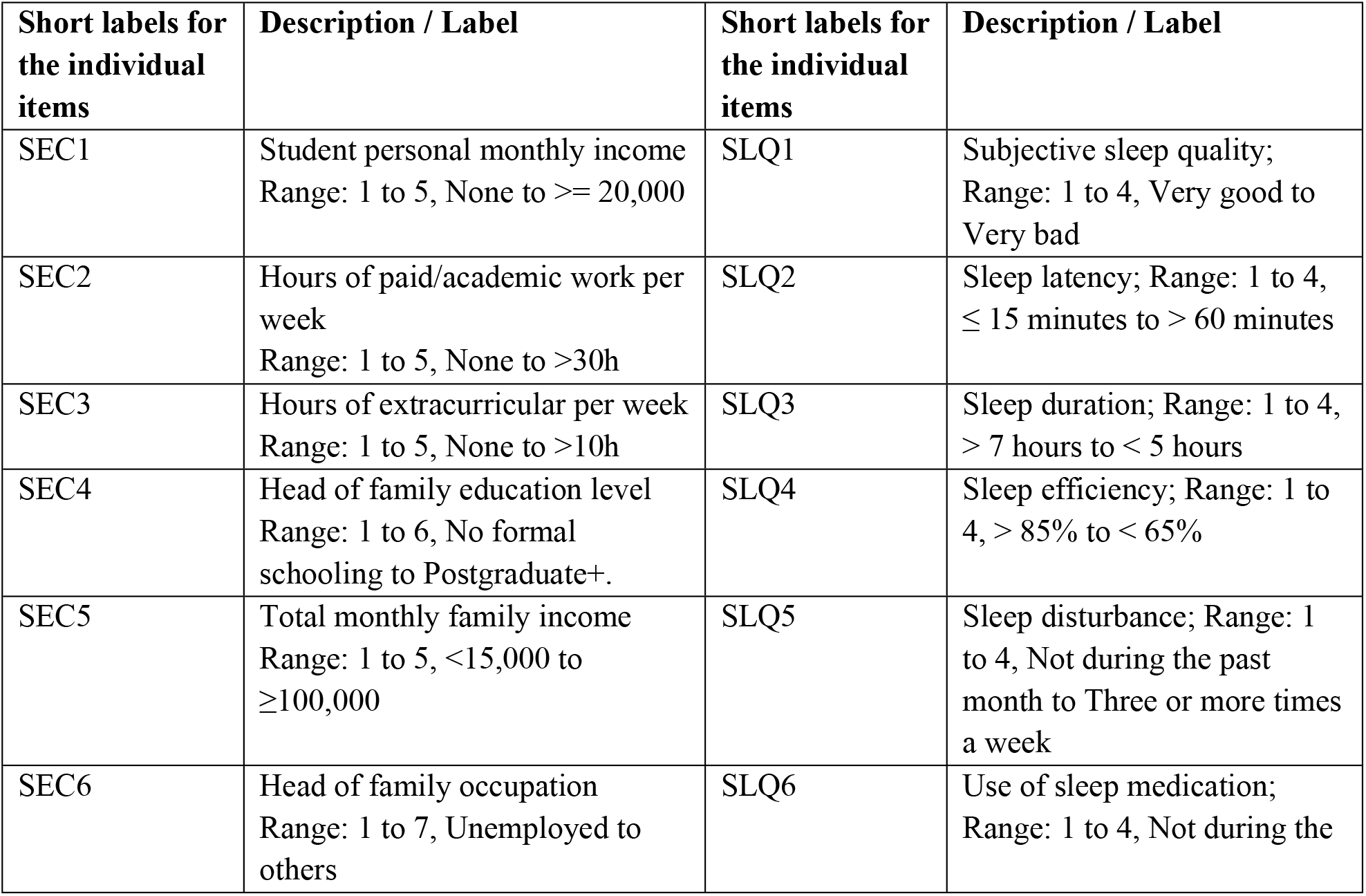
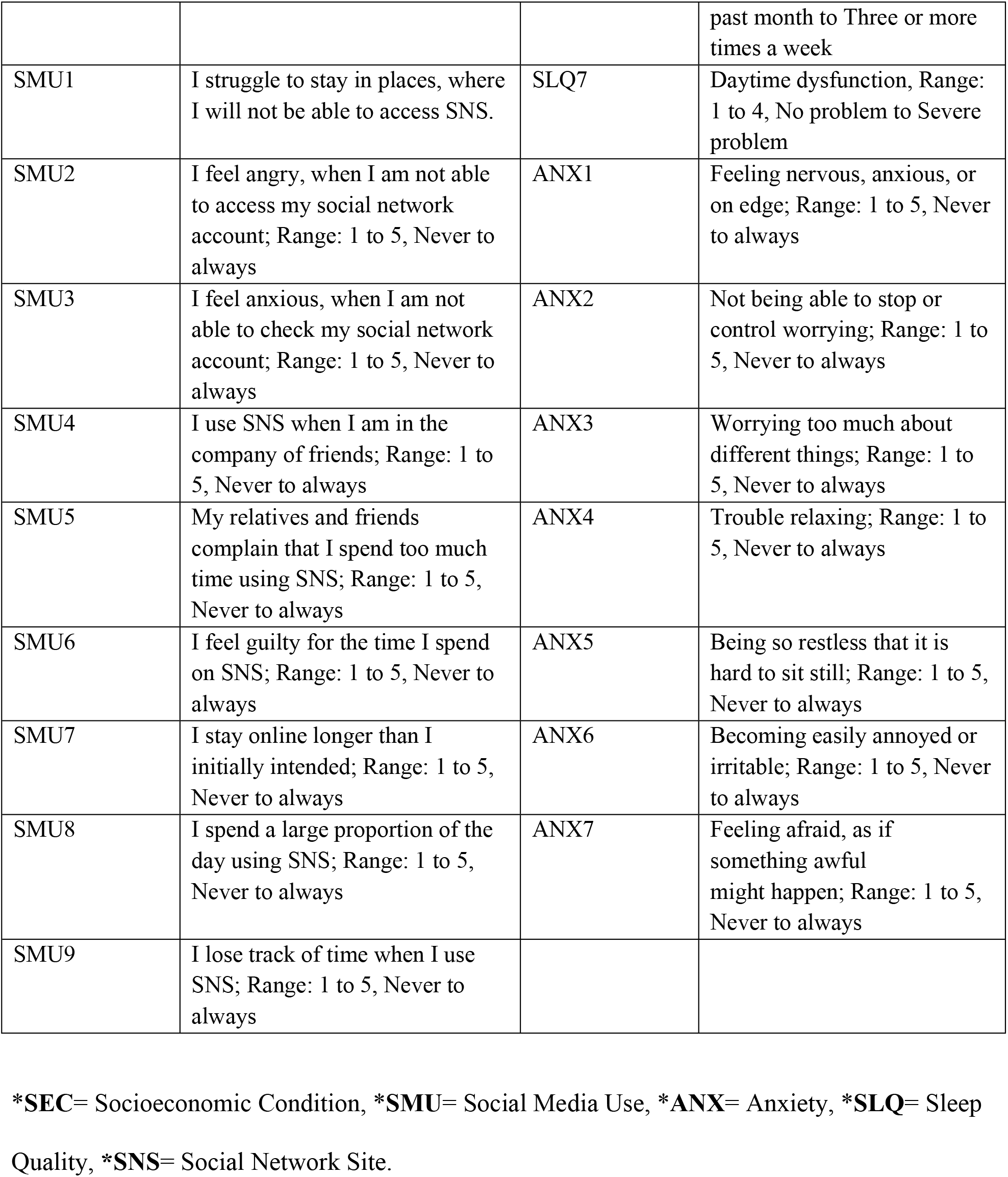
The excerpts of the questionnaire used in the research.

| Short labels for the individual items | Description / Label | Short labels for the individual items | Description / Label |
| --- | --- | --- | --- |
| SEC1 | Student personal monthly income<br>Range: 1 to 5, None to $\geq 20,000$ | SLQ1 | Subjective sleep quality;<br>Range: 1 to 4, Very good to Very bad |
| SEC2 | Hours of paid/academic work per week<br>Range: 1 to 5, None to $>30h$ | SLQ2 | Sleep latency; Range: 1 to 4, $\leq 15$ minutes to $> 60$ minutes |
| SEC3 | Hours of extracurricular per week<br>Range: 1 to 5, None to $>10h$ | SLQ3 | Sleep duration; Range: 1 to 4, $> 7$ hours to $< 5$ hours |
| SEC4 | Head of family education level<br>Range: 1 to 6, No formal schooling to Postgraduate+. | SLQ4 | Sleep efficiency; Range: 1 to 4, $> 85\%$ to $< 65\%$ |
| SEC5 | Total monthly family income<br>Range: 1 to 5, $<15,000$ to $\geq 100,000$ | SLQ5 | Sleep disturbance; Range: 1 to 4, Not during the past month to Three or more times a week |
| SEC6 | Head of family occupation<br>Range: 1 to 7, Unemployed to others | SLQ6 | Use of sleep medication;<br>Range: 1 to 4, Not during the past month to Three or more times a week |
| SMU1 | I struggle to stay in places, where I will not be able to access SNS. | SLQ7 | Daytime dysfunction, Range: 1 to 4, No problem to Severe problem |
| SMU2 | I feel angry, when I am not able to access my social network account; Range: 1 to 5, Never to always | ANX1 | Feeling nervous, anxious, or on edge; Range: 1 to 5, Never to always |
| SMU3 | I feel anxious, when I am not able to check my social network account; Range: 1 to 5, Never to always | ANX2 | Not being able to stop or control worrying; Range: 1 to 5, Never to always |
| SMU4 | I use SNS when I am in the company of friends; Range: 1 to 5, Never to always | ANX3 | Worrying too much about different things; Range: 1 to 5, Never to always |
| SMU5 | My relatives and friends complain that I spend too much time using SNS; Range: 1 to 5, Never to always | ANX4 | Trouble relaxing; Range: 1 to 5, Never to always |
| SMU6 | I feel guilty for the time I spend on SNS; Range: 1 to 5, Never to always | ANX5 | Being so restless that it is hard to sit still; Range: 1 to 5, Never to always |
| SMU7 | I stay online longer than I initially intended; Range: 1 to 5, Never to always | ANX6 | Becoming easily annoyed or irritable; Range: 1 to 5, Never to always |
| SMU8 | I spend a large proportion of the day using SNS; Range: 1 to 5, Never to always | ANX7 | Feeling afraid, as if something awful might happen; Range: 1 to 5, Never to always |
| SMU9 | I lose track of time when I use SNS; Range: 1 to 5, Never to always |  |  |

\*SEC= Socioeconomic Condition, \*SMU= Social Media Use, \*ANX= Anxiety, \*SLQ= Sleep
Quality, \*SNS= Social Network Site.

### Ethics Statement

The study was conducted in accordance with ethical guidelines approved by the university ethics committee. The approval number is BBEC,JU/M 2024/08(132) and the date of approval is August 08, 2024. All participants were provided with detailed information about the study, and consent was obtained prior to the data collection process.

### Data Analysis

All the descriptive and inferential statistics were calculated using SPSS, Excel, and SPSS AMOS for Windows. The data collected from the questionnaire were coded correctly, checked for missing values and outliers, and were found to be suitable for analysis.

### Structural Equation Modeling

The structural equation model (SEM) is a statistical method with two main parts: i) the measurement model and ii) the structural model. The measurement model shows the relationship between the hidden (latent) variable and the observed variable. The structural model is used to analyze the loadings and estimate indicators. The process of building an SEM involves the following steps.

### Specify the structural model

This model is expanded to incorporate structural relationships between latent variables. It defines the paths or regression coefficients between latent variables, includes any direct effects on observed variables, and accounts for error terms in the observed variables.

#### Estimate model parameters

SEM analysis is conducted to estimate the model’s parameters. This includes determining factor loadings, regression coefficients, variances, covariances, and other parameters based on the defined model.

#### Access model fit

The fit of the SEM model to the data is assessed using fit indices. Common indices include Chi-square (χ²), comparative fit index (CFI), incremental fit index (IFI), Tucker Lewis index (TLI), Parsimonious goodness of fit index (PGFI), normal fit index (NFI), Parsimonious normed fit index (PNFI), Parsimonious comparative fit index (PCFI) and root mean square error (RMSEA). The model’s goodness-of-fit is determined by comparing it to established thresholds or benchmarks.

## Results and Discussions

### Univariate Analysis

Table 2 shows the demographic profile of the study population. A total of 225 students participated in this study. The group was mostly male (59.6%), with 40.4% female. The study found fewer than 20% of the respondents do not earn, while the remaining respondents earned in varying levels.

**Table 2:**
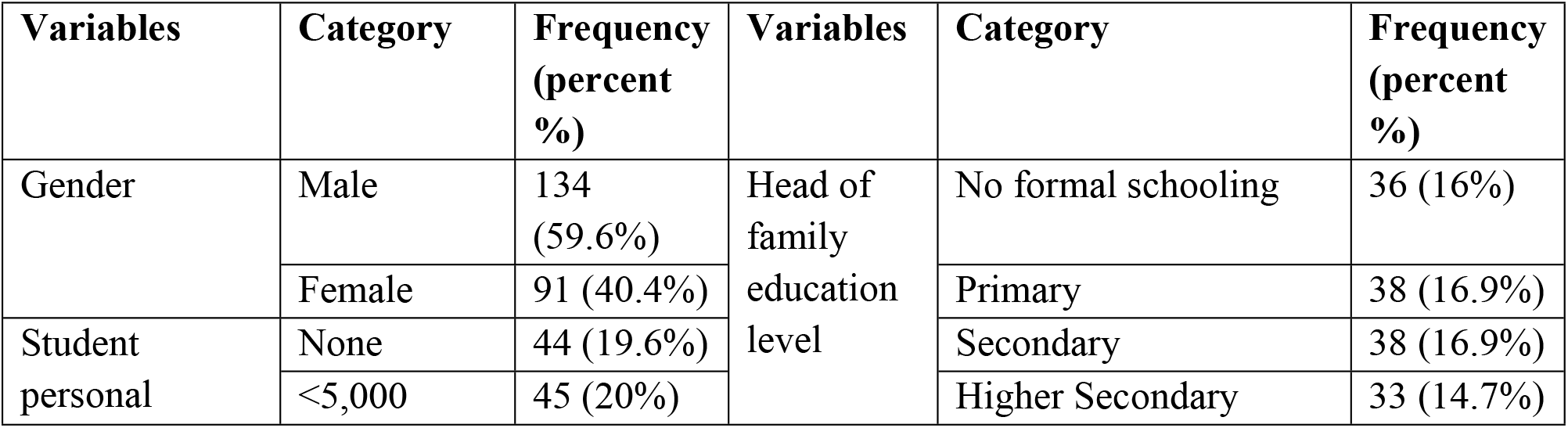
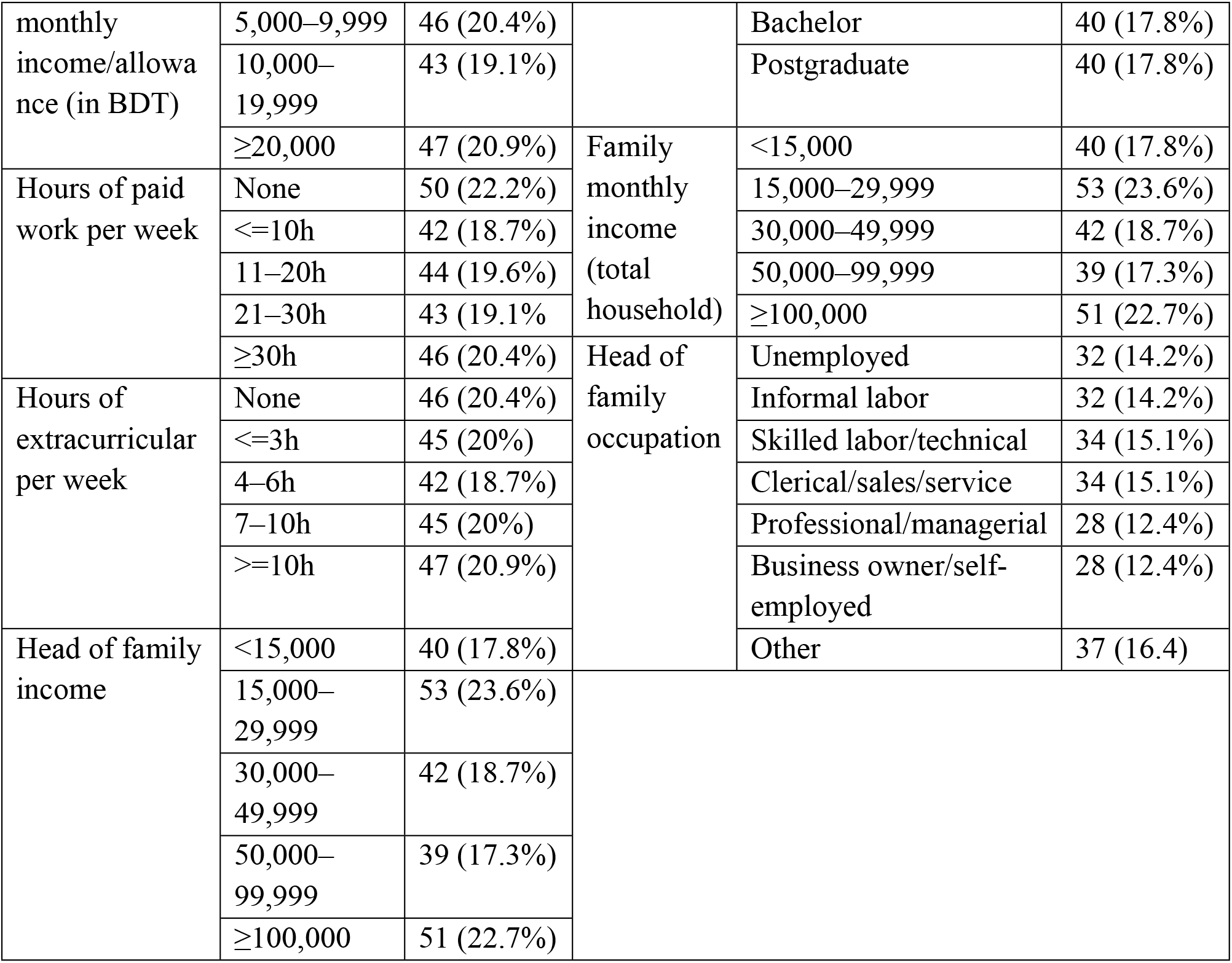
Demographic attributes of the respondents.

The subjects exhibited a relatively balanced distribution of weekly paid working hours. Approximately, 20.4% of the respondents were working 30 or more hours per week while 22.2% reported they had no working hours. Their activities outside of class varied as well. About 21% were very involved in extracurricular activities (10+ hours a week), while 20% weren’t involved at all. The respondents came from a diverse family background as well. For instance, while 18% had a parent with a bachelor’s degree, 16% had a parent with no formal schooling. Family incomes ranged widely, from moderate to relatively high.

Overall, this group of students is highly diverse in terms of their financial situation, employment, and family backgrounds.

### Bivariate Analysis

**Fig 1:**
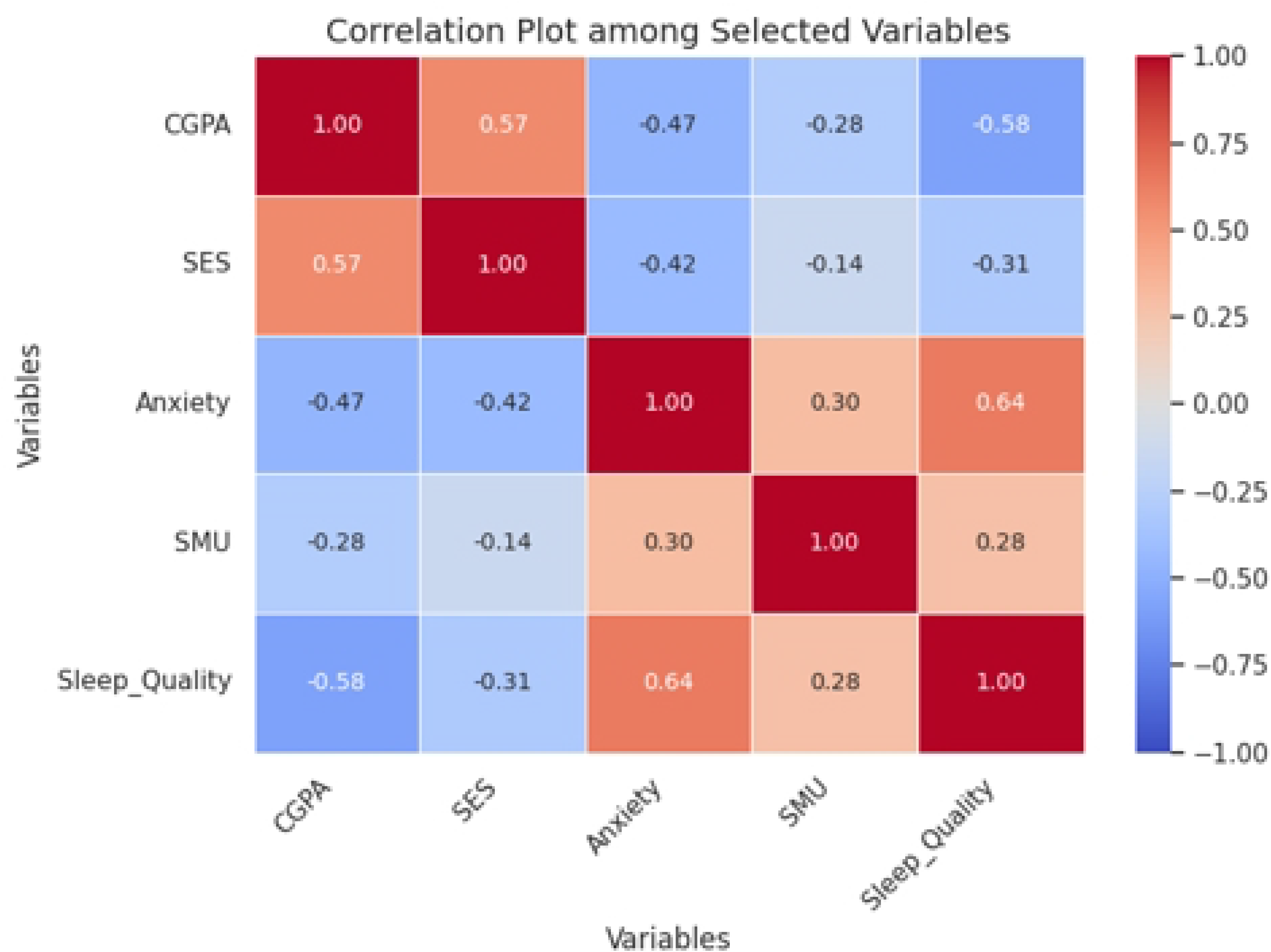
Correlation plot among selected variables.

The correlation analysis reveals several key relationships between the composite variables and academic performance (CGPA) in Fig 1. Socioeconomic score shows a positive correlation with CGPA, indicating that students from higher socioeconomic backgrounds tend to perform better academically. Anxiety has a negative correlation with CGPA, suggesting that higher anxiety levels are associated with lower academic performance. Sleep quality also exhibits a strong negative correlation with CGPA, highlighting the detrimental effect of poor sleep on academic achievement. Conversely, social media use shows a weak negative correlation with CGPA, implying that while excessive social media use slightly impacts academic performance, the effect is less significant compared to other factors.

**Fig 2:**
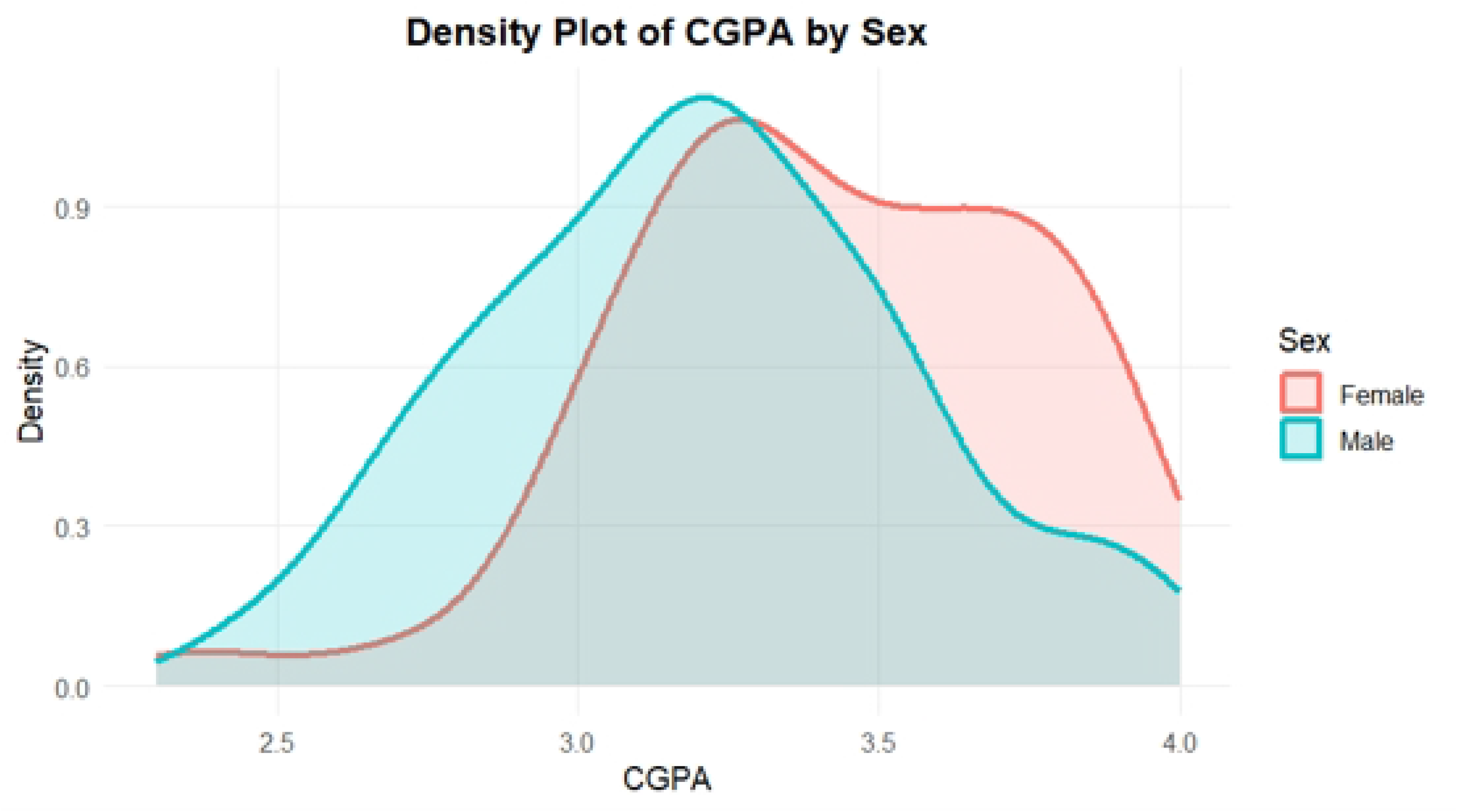
Density plot of CGPA by sex.

The density plot (Fig 2) of CGPA by sex indicates a slight difference in academic performance between male and female students. Female students appear to be more concentrated in the higher CGPA range, particularly above 3.4, whereas male students are more concentrated in the lower to middle CGPA range. Although the two distributions overlap substantially, the graph suggests that female students tend to achieve slightly better academic results overall than male students.

**Fig 3:**
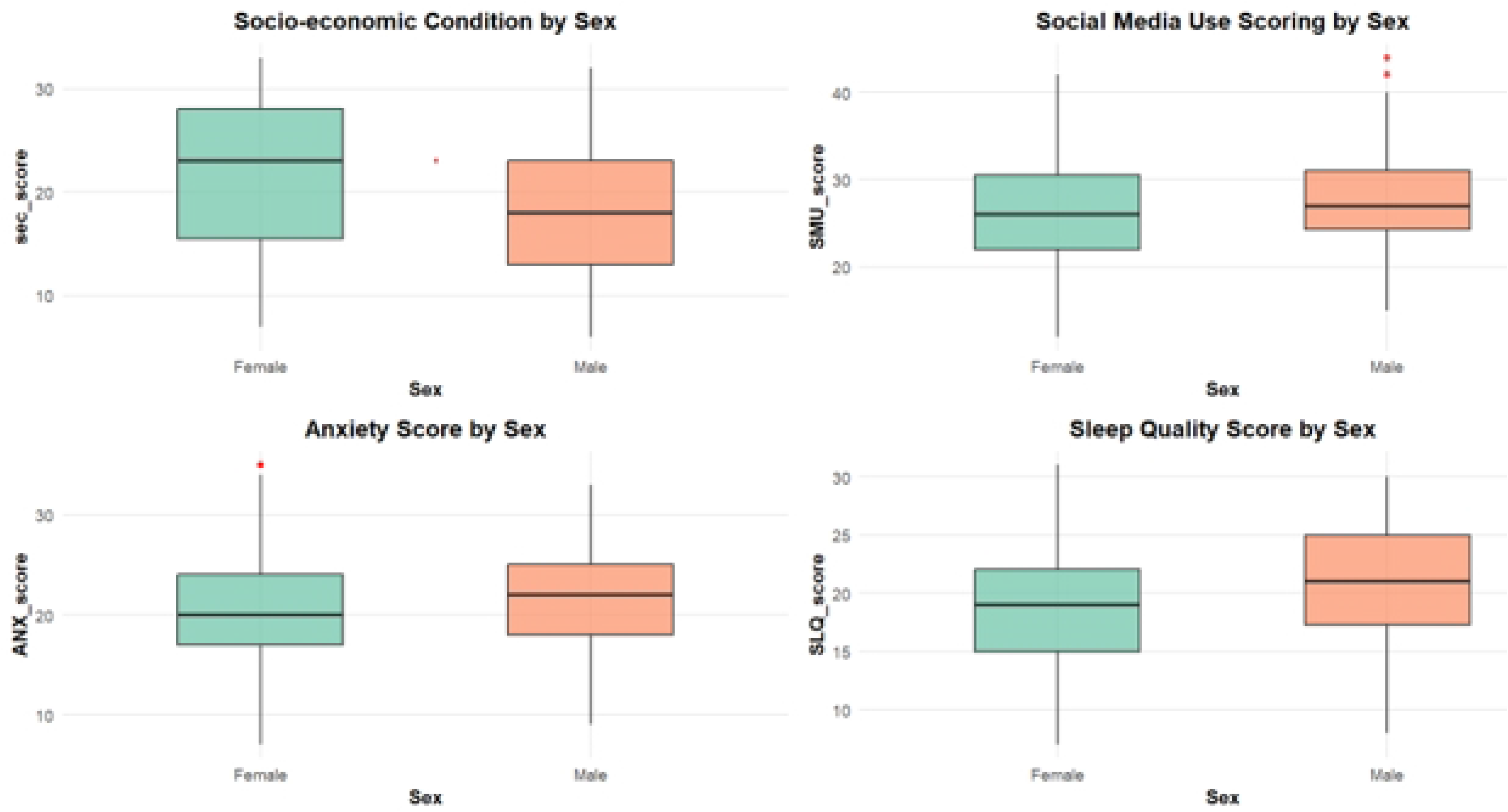
Boxplots of socioeconomic condition, social media use, anxiety, and sleep quality scores by sex.

The boxplots (Fig 3) indicate modest sex-based differences across the four variables. Female students appear to have a higher median socioeconomic score than male students, suggesting comparatively better socio-economic condition. On the other hand, male students show slightly higher median scores for social media use, anxiety, and sleep quality, although the distributions for males and females overlap considerably in all cases. This overlap suggests that the differences between the two groups are present but not very large. In addition, a few higher outliers are visible among male students in social media use. Overall, the figure suggests moderate variation by sex rather than a strong separation between male and female students.

**Table 3:**
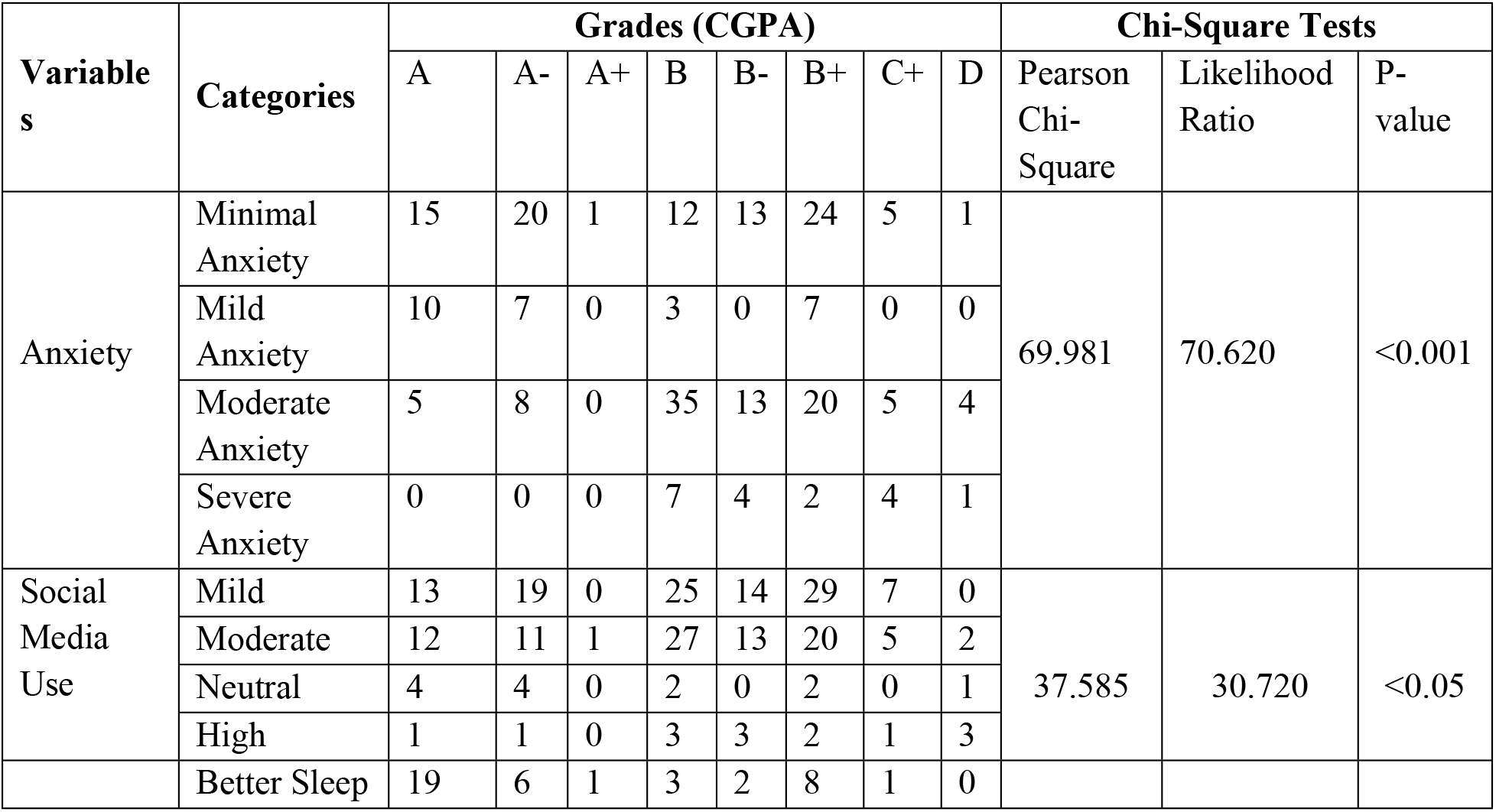
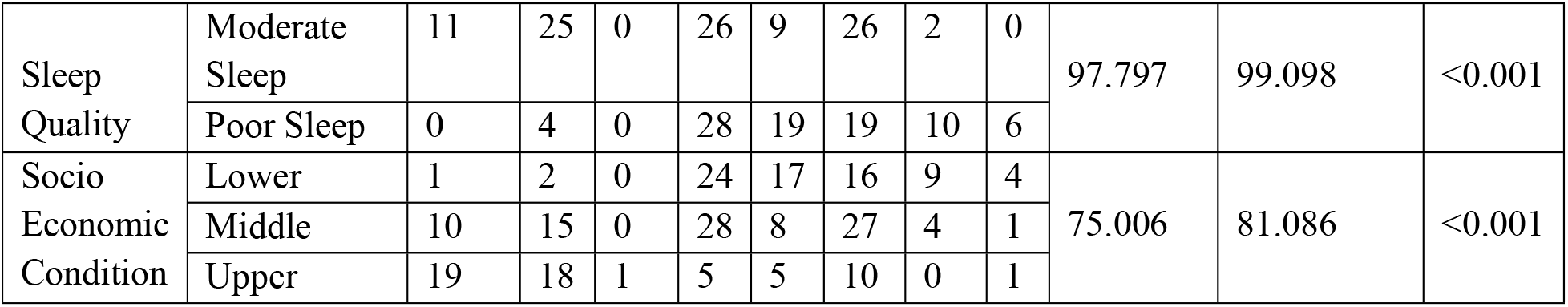
Bivariate association between CGPA grades and anxiety, social media use, sleep quality, and socioeconomic condition.

Table 3 shows that CGPA is significantly associated with anxiety, social media use, sleep quality, and socioeconomic status. Students with minimal or mild anxiety are more concentrated in the higher-grade categories, while those with moderate or severe anxiety appear more often in the lower grades. A similar pattern is observed for sleep quality, where students with better sleep are more represented in higher CGPA groups, while poor sleepers are concentrated in the lower and middle grade ranges. Socioeconomic condition also shows a significant association with grades. Students having upper socioeconomic condition are more likely to pursue a better grade than the students coming from lower and middle socio demographic background. The association between social media use and grades of the subjects is also statistically significant at 5% level of significance.

### Multivariate Analysis

#### Hypothesis statement

The hypotheses for the study are as follows, *H*_1_: Socioeconomic condition (SEC) has a significant positive effect on academic performance (CGPA), *H*_2_: Anxiety (ANX) has a significant negative effect on academic performance (CGPA), *H*_3_: Sleep quality (SQ) has a significant positive effect on academic performance (CGPA), *H*_4_: Social media use (SMU) has a significant negative effect on academic performance (CGPA).

#### Measurement model

To investigate the significant factors that may impact a student’s academic performance, a measurement model (Fig 4) has been developed, including 4 latent constructs. CFA model showed that the four constructs in this research are Socioeconomic status, Anxiety, Social Media Use, and Sleep Quality. The observed (independent) variables are represented by SEC1, SEC4-SEC6, ANX1-ANX7, SMU3, SMU8-SMU9, and SLQ1-SLQ7, and their factor loadings reflect the relationships between these variables and the unobserved (latent) constructs.

**Fig 4:**
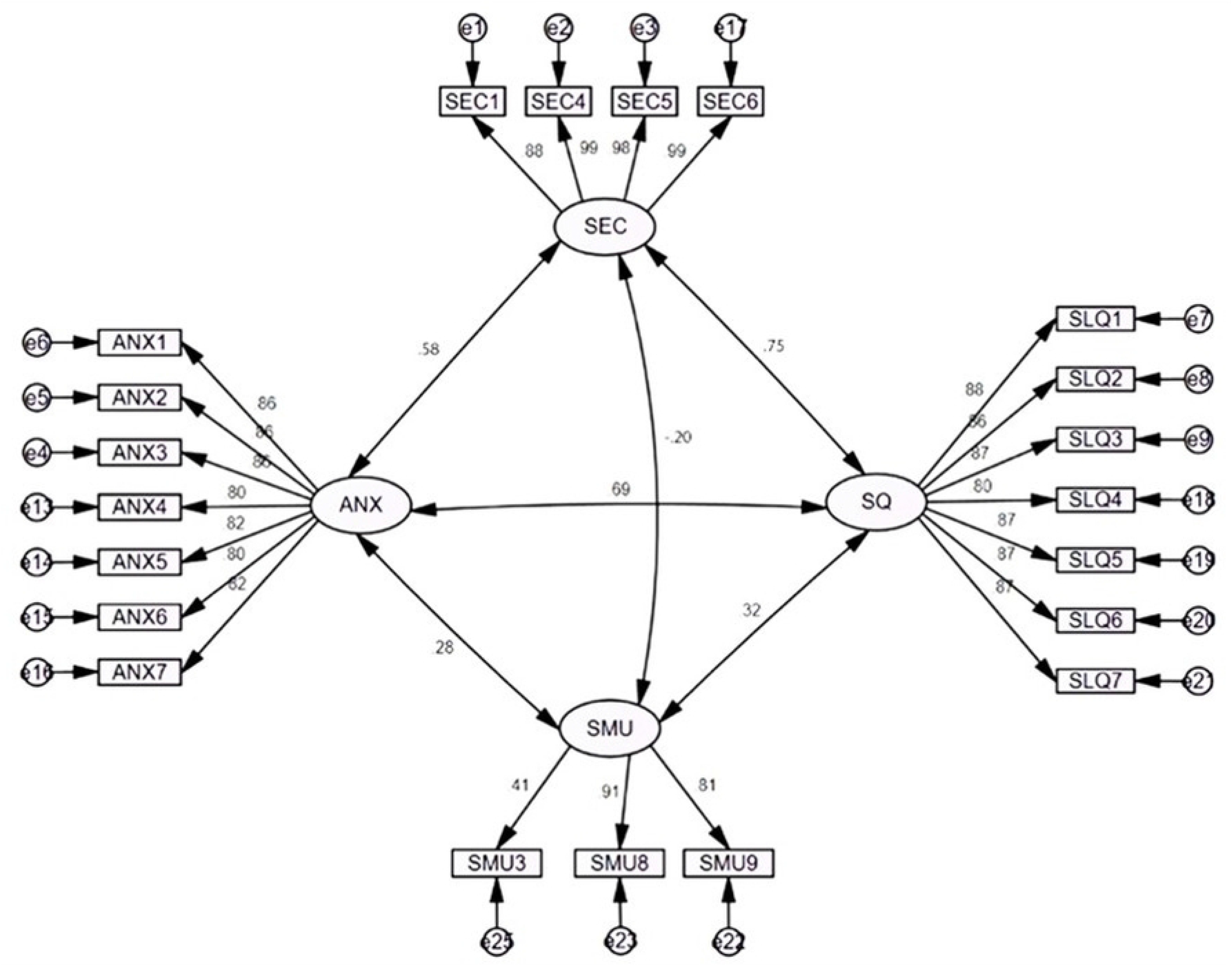
Measurement Model.

To improve the model’s reliability, validity, and better fit of the model, items with factor loadings below 0.5 were removed, except for SMU3, which had a loading of 0.414. This item has been kept because it adds valuable insight to capture the social media use construct [22, 23]. Most of the factor loadings value range from 0.80 to 0.99, suggesting that the observed variables are strongly associated with the latent variables of socioeconomic condition (SEC), anxiety (ANX), sleep quality (SQ), and social media use (SMU).

#### Structural Model

To assess whether the factor structure can be replicated in the dataset of 225 participants, a structural equation model (Fig 5) is conducted following the fitting of the CFA. While there are numerous fit indices available to evaluate the goodness of fit in CFA, only a select few, as listed in Table 4, have been used in our research.

**Fig 5:**
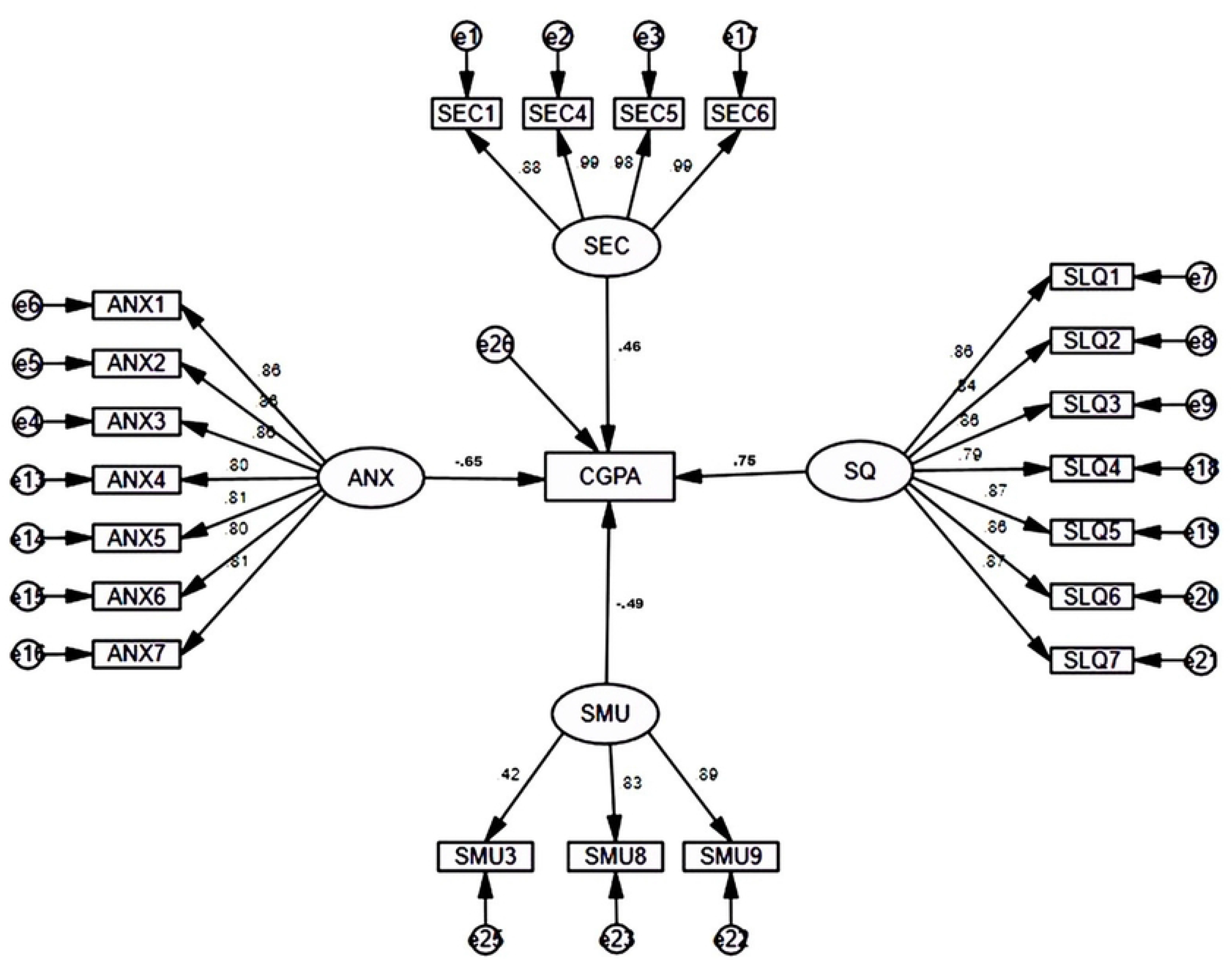
Structural Equation Model.

#### Reliability and Validity

The overall goodness of fit indices for the model has been assessed to determine how well the data fit the proposed model.

**Table 3:** Test of Reliability and Validity.

| Contrast | Item | Loading | CR (>0.70) | AVE (>0.50) | Cronbach's Alpha (>0.70) |
| --- | --- | --- | --- | --- | --- |
| Socioeconomic Condition | SEC1 | 0.878 | 0.98 | 0.92 | 0.97 |
|  | SEC4 | 0.987 |  |  |  |
|  | SEC5 | 0.981 |  |  |  |
|  | SEC6 | 0.991 |  |  |  |
| Social Media Uses | SMU3 | 0.414 | 0.75 | 0.55 | 0.71 |
|  | SMU8 | 0.908 |  |  |  |
|  | SMU9 | 0.812 |  |  |  |
| Anxiety | ANX1 | 0.862 | 0.95 | 0.69 | 0.95 |
|  | ANX2 | 0.858 |  |  |  |
|  | ANX3 | 0.855 |  |  |  |
|  | ANX4 | 0.805 |  |  |  |
|  | ANX5 | 0.817 |  |  |  |
|  | ANX6 | 0.799 |  |  |  |
|  | ANX7 | 0.818 |  |  |  |
| Sleep Quality | SLQ1 | 0.87 | 0.96 | 0.74 | 0.96 |
|  | SLQ2 | 0.860 |  |  |  |
|  | SLQ3 | 0.874 |  |  |  |
|  | SLQ4 | 0.804 |  |  |  |
|  | SLQ5 | 0.875 |  |  |  |
|  | SLQ6 | 0.871 |  |  |  |
|  | SLQ7 | 0.868 |  |  |  |

\*CR= Composite Reliability, \*AVE= Average Variance Extracted

In testing constructs’ reliability and validity, the composite reliability (CR), average variance extracted (AVE), and Cronbach’s coefficient alpha are calculated. In this study, CR values for the constructs range from 0.75 to 0.98, which exceed our minimum cut-off value of 0.70. The AVE values are greater than 0.50, and the Cronbach’s alpha score are greater than 0.70. Finally, as can be seen in table 3, all the construct with their respective indicator of our study are observed to be reliable and valid for further analysis.

**Table 4:**
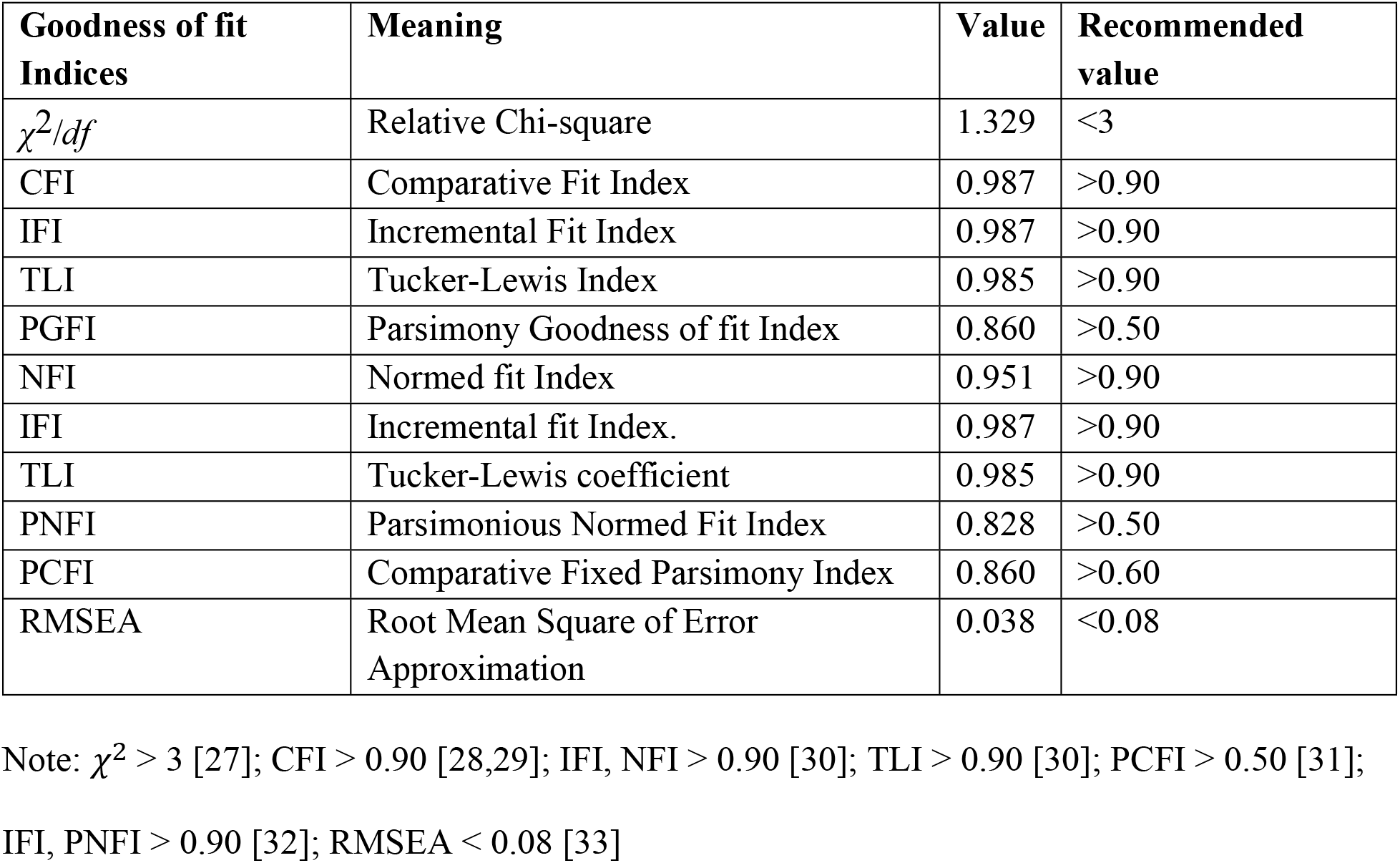
Model fit indices.

The fit indices (Table 4) show that the model fits the data well, with all values meeting or exceeding the recommended thresholds.

**Table 5:**
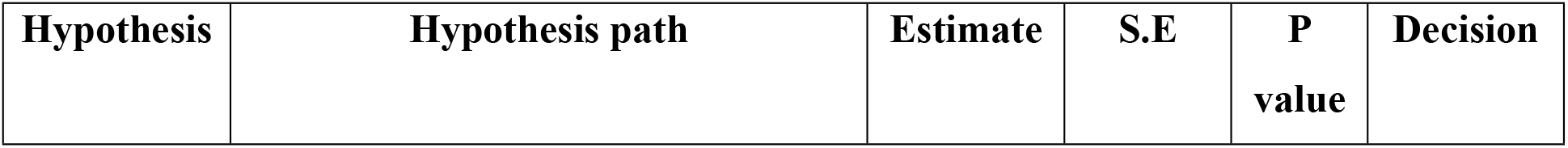
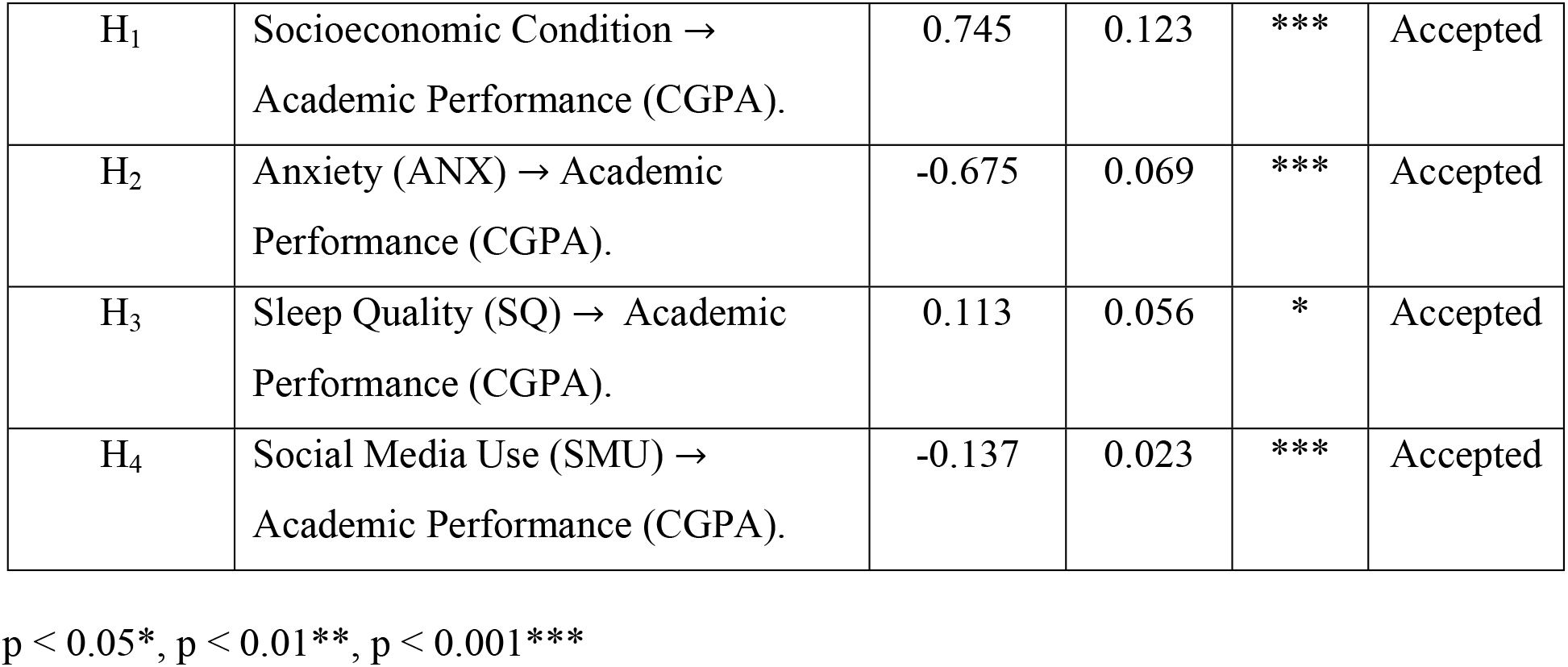
Results of hypothesis test based on SEM model.

| Hypothesis | Hypothesis path | Estimate | S.E | P value | Decision |
| --- | --- | --- | --- | --- | --- |
| H<sub>1</sub> | Socioeconomic Condition → Academic Performance (CGPA). | 0.745 | 0.123 | *** | Accepted |
| H<sub>2</sub> | Anxiety (ANX) → Academic Performance (CGPA). | -0.675 | 0.069 | *** | Accepted |
| H<sub>3</sub> | Sleep Quality (SQ) → Academic Performance (CGPA). | 0.113 | 0.056 | * | Accepted |
| H<sub>4</sub> | Social Media Use (SMU) → Academic Performance (CGPA). | -0.137 | 0.023 | *** | Accepted |

$p < 0.05^*$ , $p < 0.01^{**}$ , $p < 0.001^{***}$

Research hypotheses were investigated based on path coefficients and showed in table 5. Firstly, hypothesis H_1_ (socioeconomic condition is positively related to academic performance) showed a strong positive relationship (β = 0.745, p < 0.001), indicating that higher socioeconomic status is associated with better academic performance. Secondly, hypothesis H_2_ (anxiety is negatively related to academic performance) also demonstrated a negative relationship with an acceptable strength (β = -0.675, p < 0.001). Thirdly, hypothesis H_3_ (sleep quality is positively related to academic performance) revealed a significant positive association (β = 0.113, p < 0.05), indicating that better sleep quality is associated with better academic performance. Finally, hypothesis H_4_ (social media use (SMU) is related to academic performance) showed a negative relation with acceptable strength (β = -0.137, p < 0.001), which shows social media usage have a significant reciprocal effect on academic performance of students.

## Discussion & Conclusion

This study explores the interplay of anxiety, sleep quality, social media use, and socioeconomic status on academic performance among Bangladeshi public university students. The findings not only align with previous research but also add new insights into how these four factors collectively affect CGPA. By employing Structural Equation Modeling (SEM), the study provided a comprehensive analysis of these factors and their complex interrelationships, contributing valuable insights to the literature on student well-being and academic success.

The first hypothesis about educational progress and socioeconomic backgrounds of the students is strongly supported by the data, as SEM results reveal a significant positive relationship, indicating that students from higher socioeconomic backgrounds tend to perform better academically, which aligns with previous research [34]. Additionally, female students outperformed male students, supporting [35], which indicates that girls may benefit more from higher social economic background, as the gender achievement gap shrinks in higher socioeconomic groups [36]. In this study, female students had higher monthly income and family income than male students, suggesting they may receive more financial support, which could improve access to educational resources. Students with higher SEC showed better academic achievement in all cycles analyzed [37], consistent with other studies emphasizing the role of socio-economic opportunities on academic performance [13,38–40]. As students’ financial resources grow, factors like family support, tutoring, and access to additional resources play a key role in enhancing performance [41–44]. Conversely, students from lower societal backgrounds face greater challenges and systemic barriers to academic success [45–47]. This study also found a robust negative relationship between social anxiety and results of the pupils reflecting that higher anxiety levels have a profound impact on academic performance. This finding also supports previous research conducted in the same notion [48–50]. This study also shows a slight difference in anxiety levels according to gender. While anxiety affects both genders, females tend to report slightly lower levels of anxiety across several items compared to male. The analysis further reveals that a significant number of male candidates come from lower socioeconomic backgrounds. This could be one contributing factor to the higher levels of anxiety observed among males, as they often face financial crises, which exacerbate their stress. The constant struggle to meet financial needs and overcome economic hardships may increase the overall anxiety levels in males, as they are more likely to experience the pressures of financial instability. This financial strain, combined with the added responsibilities they may carry, can significantly amplify their anxiety.

The third hypothesis suggests a positive relationship of sleep quality with academic performance. This can be attributed to the smaller effect size compared to the other variables. It indicates that good sleep helps students concentrate in class and remain stress-free, contributing to better performance. These findings align with previous research, which demonstrated that adequate sleep enhances academic performance [12,51–52]. However, the exact nature of this relationship can vary depending on other influencing factors [53–54]. Poor sleep quality negatively impacts cognitive function, leading to difficulties in concentration, memory retention, and overall mental clarity, which ultimately hinders academic achievement. This underscores the critical role of both sleep quantity and quality in supporting students’ academic success. The fourth hypothesis about the association between educational progress and social media use found to be significantly associated resulting negative path coefficient which alligns with previous studies showing a substantial negative effect on Bangladeshi students’ performance [55–58]. This might be because excessive use of social media platforms can shrink the study hour and disrupt the attention and focus, which can also lead to procrastination and less academic engagement among students. Given its widespread use for academic purposes, social media’s impact may not always be harmful, depending on how students engage with it. This opens new avenues for exploring the nuanced effects of social media on academic performance.

This study provides valuable insights into the complex interplay of anxiety, sleep quality, social media use, and socioeconomic status on the academic performance of Bangladeshi public university students. The findings confirm that excessive use of online social media platforms can have a negative impact on students’ educational progress. This study also observed students from higher socioeconomic backgrounds and those with lower anxiety levels tend to perform better academically. While good sleep quality positively influences academic success, its effect is not as strong as that of anxiety or socioeconomic status. Overall, this study contributes to the growing body of literature on student well-being and academic achievement, offering new directions for future research.

### Limitation of the study

One key limitation of this study is the use of a non-parametric convenience sampling technique, which may result in sampling bias. The sample was drawn from a limited number of public universities in Bangladesh, and participants were self-selected. This could have led to a skewed representation of the student population, as students with certain characteristics or experiences may have been more inclined to participate, thus limiting the generalizability of the findings to all university students in Bangladesh. Additionally, the reliance on self-reported data introduces potential response biases. Since participants were asked to report their own levels of anxiety, sleep quality, and social media use, there is a risk of inaccuracies due to social desirability or recall bias, with students possibly underreporting negative behaviors such as excessive social media use or poor sleep quality.

Furthermore, the study was conducted under time and budget constraints, which may have limited the ability to capture a broader and more diverse sample of students. The data collection occurred within a short time frame, possibly affecting the depth of information gathered, and budget limitations may have prevented the use of more extensive resources or advanced measurement tools. Despite these limitations, the study was able to meet its primary objectives and provide valuable insights into the factors influencing academic performance among Bangladeshi public university students. However, these constraints should be considered when interpreting the findings, as they may have impacted the scope and generalizability of the results.

## Data Availability

N/A

https://docs.google.com/spreadsheets/d/1KMv9R0VuQTXGXShxXR6Mc-Twp0PEC4F0/edit?usp=drive_link&ouid=113320289093022702205&rtpof=true&sd=true

## AI Declaration

Artificial Intelligence (ChatGPT -5.4) was used to enhance the clarity of the manuscript and assist with language editing.

